# Machine Learning based COVID-19 Diagnosis from Blood Tests with Robustness to Domain Shifts

**DOI:** 10.1101/2021.04.06.21254997

**Authors:** Theresa Roland, Carl Böck, Thomas Tschoellitsch, Alexander Maletzky, Sepp Hochreiter, Jens Meier, Günter Klambauer

## Abstract

We investigate machine learning models that identify COVID-19 positive patients and estimate the mortality risk based on routinely acquired blood tests in a hospital setting. However, during pandemics or new outbreaks, disease and testing characteristics change, thus we face domain shifts. Domain shifts can be caused, e.g., by changes in the disease prevalence (spreading or tested population), by refined RT-PCR testing procedures (taking samples, laboratory), or by virus mutations. Therefore, machine learning models for diagnosing COVID-19 or other diseases may not be reliable and degrade in performance over time. To countermand this effect, we propose methods that first identify domain shifts and then reverse their negative effects on the model performance. Frequent re-training and reassessment, as well as stronger weighting of more recent samples, keeps model performance and credibility at a high level over time. Our diagnosis models are constructed and tested on large-scale data sets, steadily adapt to observed domain shifts, and maintain high ROC AUC values along pandemics.

## 1 Introduction

Reverse transcription polymerase chain reaction (RT-PCR)1 are still the gold standard tests for the coronavirus disease 2019 (COVID-19)^2^. However, RT-PCR tests are expensive, time-consuming, and not suited for high-throughput or large-scale testing efforts. In contrast, antigen tests are cheap and fast, but they come with considerably lower sensitivity than RT-PCR tests^3^. Blood tests for COVID-19 are a promising technique, since they unify the best of RT-PCR and antigen tests: they are cheap, fast, efficient, and have sufficiently high sensitivity when combined with machine learning (ML) methods. Furthermore, automatically checking all routinely taken blood tests for COVID-19 allows frequent, fast and broad scanning at low costs, thus provides a powerful tool to ban new outbreaks^4,5^. Therefore, we assess ML methods for diagnosing COVID-19 from blood tests. ML can enhance the sensitivity of cheap and fast tests such as antigen^6^ or blood tests, therefore enabling a cost efficient alternative to RT-PCR tests. ML methods enhanced tests could be particularly useful for asymptomatic patients with a routine blood test, who would not be tested for COVID-19. In this scenario, COVID-19 positive patients could be identified, isolated and a further spread of the virus might be prevented. Especially in developing countries with limited testing capacities, the ML enhanced tests can evolve into an efficient tool in combating a pandemic.

To confine the spread of infectious diseases, and especially the COVID-19 pandemic, ML approaches can be applied in very different ways^7^. ML algorithms help in developing vaccines and drugs for the treatment of COVID-19^8–10^. COVID-19 and the patient’s prognosis can be predicted from chest CT-scans, X-rays^11–14^ or sound recordings of coughs or breathing^15–17^. Furthermore, it has been shown that ML models based on blood tests are capable of detecting COVID-19 infection^18–32^ and predicting other outcomes, such as survival or admission to an intensive care unit^33–41^.

An ML model is constructed via learning on a data set with the goal that the model generalizes well, that is, performs well on new, unseen data, e.g., correctly predicts the label or class for a new data item. The quality, size and characteristics of the training data set strongly determine the predictive quality of the resulting model on new data. The central ML paradigm is that training data and future (test) data have the same distributions. This paradigm guarantees that the constructed or learned model generalizes well to future data and has high predictive performance on new data. However, this paradigm is violated during pandemics. Data sets collected during the progression of the COVID-19 pandemic are characterized by strong changes in distribution, called domain shifts. These domain shifts violate the central ML paradigm, nevertheless, they were insufficiently considered or even neglected during the evaluation of ML models. Unexpected behavior of the models in real world hospital settings often stem from neglected domain shifts^42^. Such an unexpected behavior could even lead to unfavorable consequences, like a major disease outbreak in a hospital. Most of the previous ML studies evaluated the predictive performance of the learned models by cross-validation, bootstrapping or fixed splits on randomly drawn samples^18–22,26–32^. However, the theoretical justification of these evaluation methods is heavily founded on the central ML paradigm: that the distributions remain constant over time. To disregard domain shifts is a culpable negligence, since they may lead to an overoptimistic performance estimate on which medical practitioners base their decisions. These decisions are then misguided.

Yang et al.^25^ and Plante et al.^24^ addressed the domain shifts via evaluation on an external data set. Yang et al.^25^ trained and evaluated their models on data from the same period and therefore, temporal domain shifts were not sufficiently considered. The training and external evaluation set as in Plante et al. ^24^ only includes pre-pandemic negatives, they missed out on using pandemic negatives. Soltan et al.^23^ considered the temporal domain shift by conducting a prospective evaluation. However, analogous to Plante et al.^24^, the negatives are all pre-pandemic, therefore, the domain shift is artificially generated and can deviate from domain shifts during the pandemic.

In the following, we describe the categories of domain shifts that can occur in COVID-19 data sets. For the categorization, we have to consider two random variables, which both are obtained by testing a patient:

- *x*: *Outcome of a fast and cheap test*. The measurement values for a patient, which serve as input data (input features) for an ML model. We assume that the COVID-19 status (positive / negative) can, to some extent, be inferred from these tests. The measurements can arise from a fast and cheap test such as a blood test or vital sign measurement. To illustrate this value, we assume that *x* is the *fibrinogen* level, since it tends to rise during a systemic inflammation^43^.
- *y*: *Outcome of the slow and expensive COVID-19 RT-PCR test*, which is assumed to be binary *y* ∈{0, 1} to indicate the COVID-19 status. The test result *y* is assumed to be the ground truth and should give the actual COVID-19 status.

Our goal is to use ML methods to predict *y* from *x*, in order to replace the slow and expensive COVID-19 RT-PCR test by a fast and cheap test.

Examples of temporal domain shifts are shown in Figure 1 **a**, which affect the model performance and the trustworthiness of performance estimates, see Figure 1 **b** and **c**. We identify and define following categories of domain shifts^44,45^:

**Figure 1:**
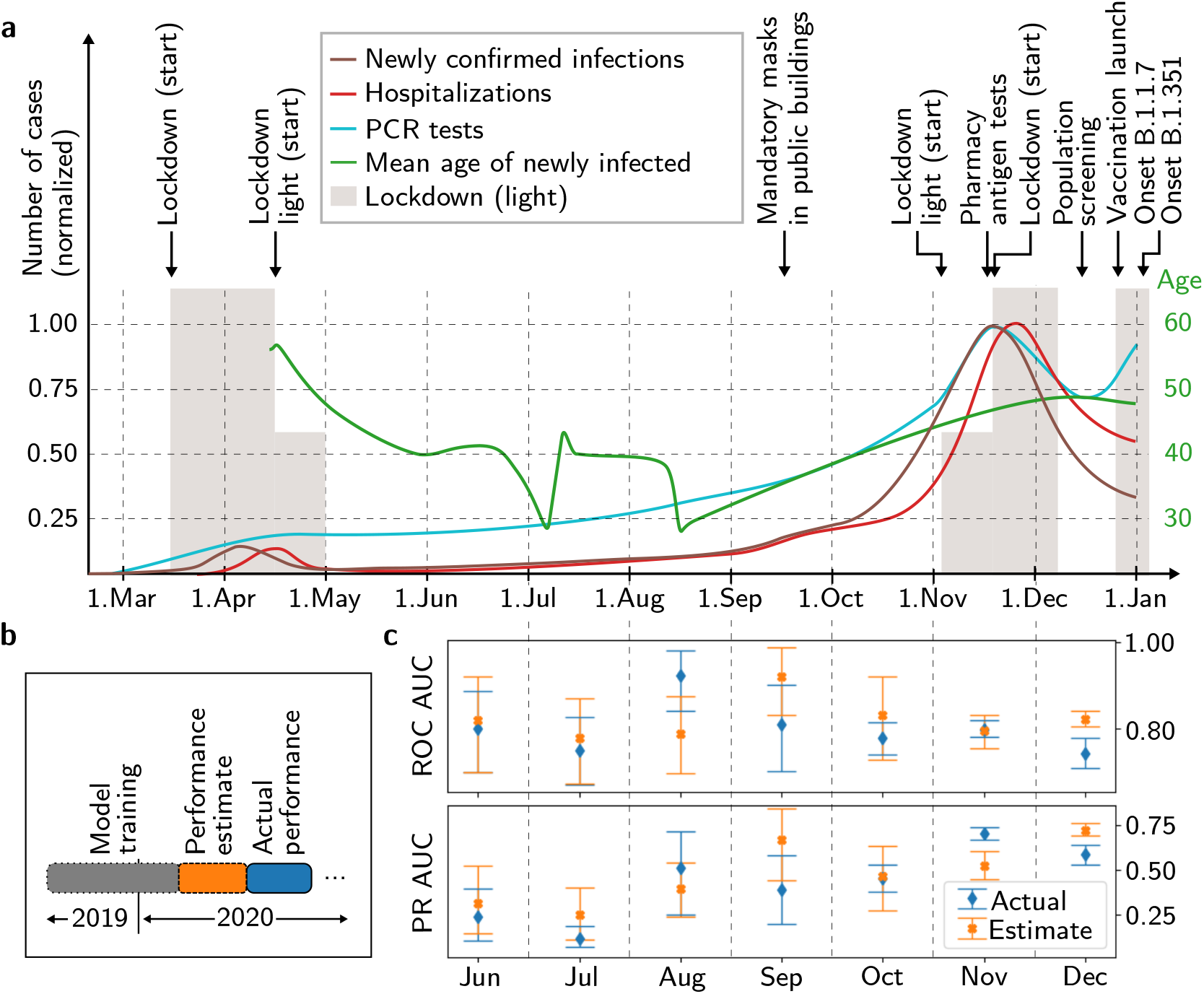
Domain shifts in COVID-19 data sets. **a**, COVID-19 numbers in Austria over time, illustrating factors causing a temporal domain shift. The numbers are sketched according to data from the Austrian BMSGPK (https://www.data.gv.at/COVID-19/). **b**, The actual model performance is calculated for each month from June to December 2020 and the estimated model performance is calculated on the respective previous month. **c**, Estimated and actual performance with 95 % confidence intervals. The estimated and actual ROC AUC is significantly different in December and PR AUC differs significantly in November and December, showing the effect of the domain shifts. Note that the PR AUC is sensitive to changes of prevalence.

- **Prior shift**: *p*(*y*). The probability of observing a certain RT-PCR test result, e.g., *y* = 1, strongly changes during the pandemic. If the overall prevalence of the disease in the population is high, the probability to observe a positive test usually increases.
- **Covariate shift**: *p*(*x*). The distribution of the patient features is also affected by the overall pandemic course. E.g., if the prevalence of the disease is high, more persons suffer from disease symptoms, with potentially high *fibrinogen*, and go to the hospital. Nevertheless, *fibrinogen* levels could also change without connection to the pandemic, for example, with time of year^46^. Or, in case there is an obligation for testing, the person group is changed as are the measurements.
- **General domain shift**: *p*(*y, x*). The joint distribution of patient features and labels also changes during the pandemic, for example with new virus mutations. A new mutation could lead to more severe disease progression^47^ and to even higher *fibrinogen*.
- **Concept shift**: *p*(*y*|*x*). Probability to observe a certain RT-PCR test result given a patient characterized by their measurements such as blood tests. We model this by *p*(*y* = 1|*x*) ≈ *g*(*x*; *w*), with the model *g* and the model parameters *w*. The RT≈PCR test result *y* changes even if the patient features *x* are the same, which can occur with changing test technologies, changing test procedures, changing thresholds, and so on.

Neglecting and insufficiently countering the above mentioned domain shifts can lead to undesired consequences and failures of the models:

### Unreliable performance estimates

Performance estimates without consideration of domain shifts might be overoptimistic and the actual performance of the model can deviate significantly from the estimate^42^ (see Figure 1, and Section 2.5).

### Degrading of predictive performance over time

Standard ML approaches are unable to cope with domain shifts over time and during the progression of a pandemic, which can result in a decrease of predictive performance^44,48,49^.

In light of the domain shifts, we suggest lifelong learning and assessment^50–52^, thereby maximizing the clinical utility of the models. Concretely, we propose a) frequent temporal validation to identify domain shifts and b) re-training the models with higher weights of recently acquired samples. To this end, a continuous stream of COVID-19 samples is required, which can be achieved by routinely testing a subset of samples with an RT-PCR test.

We evaluate and compare our proposed approach of lifelong learning and assessment against standard ML approaches on a large-scale data set. This data set comprises 127,115 samples after pre-processing and merging, which exceeds the data set size of many small scale studies^18–22,32^ by far. Our data set comprises pre-pandemic negative samples and pandemic negative and positive samples spanning over multiple different departments of the Kepler University Hospital, Linz. As opposed to studies that require additional expensive features^19,22,23^, our models use no other features than blood test, age, gender and hospital admission type. This way, the blood tests can be automatically scanned for COVID-19 in a cost-effective way without any additional temporal effort for the hospital staff.

We additionally report the predictive ability for mortality risk of the COVID-19 positive samples on the basis of the blood tests only, again with no additional expensive features^33–35,38,40,41,53,54^. Compared to previous studies^33,36,37,39^, our mortality models are trained on a large number of COVID-19 positive patients. We again take domain shifts and other potential biases into account for mortality prediction.

## 2 Results

### 2.1 Study Cohort

Our dataset comprises 125,542 negative and 1,573 positive samples for training and evaluation of the ML models for COVID-19 diagnosis. From the negatives, 116,067 have been acquired before the pandemic and 9,475 during the pandemic. The RT-PCR test sample has been collected after the blood test, with a window of 48 hours between the two tests. From the COVID-19 diagnosis data set 919 cases survived and 118 cases died with COVID-19.

For the mortality prediction, the features and samples are selected on the basis of the COVID-19 positive patients, rather than the 2019 cohort for the COVID-19 diagnosis data set. The data sets are imbalanced in both tasks, the COVID-19 diagnosis and the mortality prediction. The pre-selection of the samples and the merging is described in more detail in Section 4 and in Figure 2. In the 2019 cohort and in the 2020 cohort women and men occur about equally often (2019 cohort: 48 % men, 2020 cohort: 48 % men). However, in the positives cohort, there are more men (56 % men). The death rate of patients relative to the COVID-19 positive samples in patients with 80 years or older is 20 %, in patients younger than 80 years, it is 8 %. In our data set, men died more than twice as often as women (68 % men). In the age group below 80 years, men died even three times as often as women with COVID-19 (75 % men).

**Figure 2:**
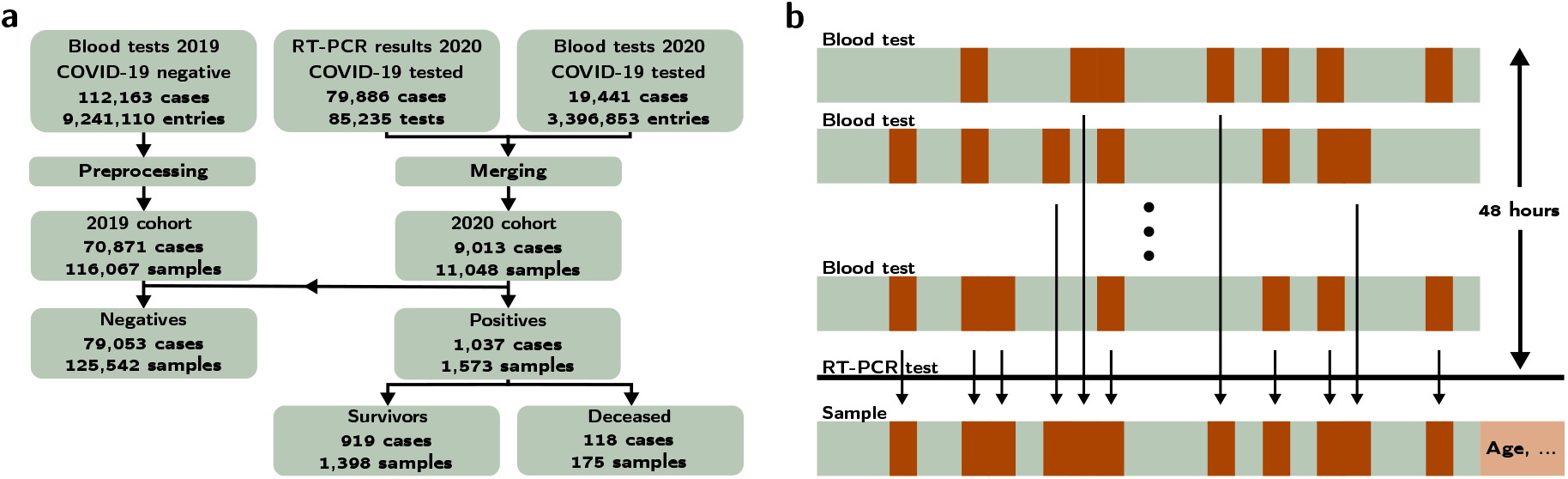
Large-scale COVID-19 data set. **a**, Block diagram of the structure of the data set. The blood tests from 2019 (blood tests 2019) are all negatives and are pre-processed to the 2019 cohort. The COVID-19 RT-PCR test results and the blood tests are merged to the 2020 cohort. The negatives data set results from the 2019 cohort and the negative samples of the 2020 cohort. The positive tested cases (positives) are further divided to the cohort with the survived and the deceased cases. Note that one case can be in the negatives and positives cohort due to a change of the COVID-19 status. Multiple samples are obtained from one case, if RT-PCR and blood tests are measured repeatedly. **b**, Aggregation of the blood tests for the COVID-19 tested patients: The blood tests of the last 48 hours before the COVID-19 test are merged to one sample. In case a feature is measured multiple times, the most recent one is inserted in the sample. Patient specific data, namely age, gender and hospital admission type, are added to the sample.

### 2.2 Machine learning methods and model selection

We show the capability of the ML models to classify COVID-19 and to predict the mortality risk. We compare the performance of self normalizing neural network (SNN)^55^, K-nearest neighbor (KNN), logistic regression (LR), support vector machine (SVM), random forest (RF) and extreme gradient boosting (XGB). XGB and RF outperform other model classes in the COVID-19 diagnosis and also in the mortality prediction. The domain shifts are exposed when comparing the evaluations on different cohorts.

The hyperparameters are selected on a validation set or via nested cross-validation to avoid a hyperparameter selection bias. Performance is estimated either via standard cross-validation or by temporal cross-validation (for details see Section 4.3).

### 2.3 Comparison of estimated and actual performance

In this experiment, we investigate the effects of a standard ML approach, in which a model is trained on data collected in a particular time-period, then assessed on a hold-out set and then deployed. Concretely, we train an XGB model on data from July 2019 until October 2020, and assess the model performance on data from November 2020. We then simulate that the model is deployed and used in December 2020. Without domain shifts, the predictive performance would remain similar, but in the presence of domain shifts, the performance would be significantly different. Thus, domain shifts are exposed by comparing actual performance with the estimated performance determined on the respective previous month, see Figure 1 **b**. The area under the receiver operating characteristic curve (ROC AUC) estimate is higher than the actual performance in most months (Figure 1 **c**). The ROC AUC performance estimate for December was significantly lower than the actual performance in December. The estimated and actual area under the precision recall curve (PR AUC) differ significantly in November and December. These results show that there is a domain shift and thus there is a necessity for up-to-date assessments, otherwise the performance estimate is not trustworthy.

### 2.4 Model performance under domain shifts

In this section, we set up five modeling experiments with two prediction tasks and different assessment strategies:

i. COVID-19 diagnosis prediction assessed by random cross-validation with pre-pandemic negatives,
ii. COVID-19 diagnosis prediction assessed by random cross-validation with recent negatives,
iii. COVID-19 diagnosis prediction assessed by temporal cross-validation,
iv. mortality prediction assessed by random cross-validation,
v. mortality prediction assessed by temporal cross-validation.

We then compare the performance estimates obtained by the assessment strategy. If the performance estimates by random cross-validation and temporal cross-validation are similar, then the underlying distribution of the data is likely to be similar over time. If the performance estimates of (ii) are different from (i), then former and current negatives follow different distributions. If performance estimates from (iii) are lower than those of (i) and (ii), the distribution of the data changes over time, hence indicating the presence of domain shifts. Equally, changing performance estimates from (iv) to (v) indicate a domain shift over time. The results in terms of threshold-independent performance metrics for the comparison of the models are shown in Table 1 **a** and **b** and in Figure 3. More information about the discriminating capability of individual features is shown in Figure 5 and in Table 5.

**Table 1:** Performance metrics. **a**, Experiment (i) - (iii) are the results of the COVID-19 diagnosis prediction. In experiment (i) the test set is randomly selected from the shuffled 2019 and 2020 cohort. In experiment (ii) the test set is a random subset of the 2020 cohort and experiment (iii) are the results of a prospective evaluation on November and December 2020. **b**, The threshold-independent metrics for mortality prediction with random shuffling of the positives set (experiment (iv)) and with prospective evaluation on November and December (experiment (v)) are listed. The ML models are trained, validated and tested with five random seeds. The mean and the standard deviation () for the ROC AUC and PR AUC are listed. **c** and **d**, Performance metrics on test set of RF for different thresholds selected on basis of the negative predictive value on the validation set (NPV val) of **c**, COVID-19 diagnosis prediction in experiment (i) and **d**, mortality prediction in experiment (iv).

| **a** Threshold-independent metrics for COVID-19 diagnosis prediction |  |  |  |  |  |  |
| --- | --- | --- | --- | --- | --- | --- |
| Model | Experiment (i) |  | Experiment (ii) |  | Experiment (iii) |  |
|  | ROC AUC | PR AUC | ROC AUC | PR AUC | ROC AUC | PR AUC |
| RE | 0.5000±0.0000 | 0.0124±0.0000 | 0.5000±0.0000 | 0.0822±0.0000 | 0.5000±0.0000 | 0.3162±0.0000 |
| BF | 0.6745±0.0000 | 0.0221±0.0000 | 0.6774±0.0000 | 0.3141±0.0000 | 0.6623±0.0000 | 0.5716±0.0000 |
| SNN | 0.9567±0.0025 | 0.4349±0.0306 | 0.8998±0.0044 | 0.5577±0.0074 | 0.7836±0.0053 | 0.6620±0.0082 |
| KNN | 0.9071±0.0000 | 0.3137±0.0000 | 0.8432±0.0000 | 0.4486±0.0000 | 0.7209±0.0000 | 0.5712±0.0000 |
| LR | 0.9600±0.0008 | 0.4126±0.0145 | 0.8878±0.0022 | 0.4770±0.0086 | 0.7732±0.0008 | 0.6467±0.0059 |
| SVM | 0.9611±0.0000 | 0.4268±0.0000 | 0.9045±0.0000 | 0.5573±0.0000 | 0.7759±0.0000 | 0.6387±0.0000 |
| RF | **0.9654±0.0005** | 0.5231±0.0106 | 0.9138±0.0025 | 0.5761±0.0100 | 0.7957±0.0025 | 0.6626±0.0049 |
| XGB | 0.9629±0.0000 | **0.5558±0.0000** | **0.9169±0.0000** | **0.6216±0.0000** | **0.8142±0.0000** | **0.7077±0.0000** |

| **b** Threshold-independent metrics for mortality prediction |  |  |  |  |
| --- | --- | --- | --- | --- |
| Model | Experiment (iv) |  | Experiment (v) |  |
|  | ROC AUC | PR AUC | ROC AUC | PR AUC |
| RE | 0.5000±0.0000 | 0.1592±0.0351 | 0.5000±0.0000 | 0.1320±0.0000 |
| BF | 0.7599±0.0748 | 0.4320±0.1021 | 0.7483±0.0000 | 0.3938±0.0000 |
| SNN | 0.8656±0.0356 | 0.5866±0.1196 | 0.8478±0.0053 | 0.4917±0.0110 |
| KNN | 0.8207±0.0550 | 0.5527±0.1137 | 0.8272±0.0000 | 0.4669±0.0000 |
| LR | 0.8613±0.0351 | 0.5555±0.1281 | 0.8388±0.0088 | 0.4784±0.0173 |
| SVM | 0.8587±0.0306 | 0.5679±0.1010 | 0.8271±0.0000 | 0.4185±0.0001 |
| RF | **0.8813±0.0214** | **0.6267±0.1065** | **0.8572±0.0071** | **0.5556±0.0127** |
| XGB | 0.8501±0.0210 | 0.5196±0.1005 | 0.8038±0.0000 | 0.4334±0.0013 |

| **c** Threshold-dependent metrics of RF in experiment (i) |  |  |  |  |
| --- | --- | --- | --- | --- |
| NPV val | 0.999 | 0.995 | 0.990 | 0.980 |
| NPV | 0.999±0.000 | 0.995±0.000 | 0.990±0.000 | 0.988±0.000 |
| PPV | 0.066±0.002 | 0.414±0.015 | 0.823±0.014 | 1.000±0.000 |
| BACC | 0.887±0.002 | 0.812±0.005 | 0.588±0.003 | 0.501±0.001 |
| ACC | 0.834±0.007 | 0.984±0.000 | 0.989±0.000 | 0.988±0.000 |
| Sensitivity | 0.941±0.004 | 0.635±0.010 | 0.176±0.006 | 0.002±0.003 |
| Specificity | 0.832±0.007 | 0.989±0.000 | 1.000±0.000 | 1.000±0.000 |
| F1 | 0.124±0.004 | 0.501±0.009 | 0.290±0.008 | 0.004±0.006 |
| Threshold | 0.081±0.040 | 0.444±0.098 | 0.931±0.020 | 0.995±0.001 |

| **d** Threshold-dependent metrics of RF in experiment (iv) |  |  |  |  |  |  |
| --- | --- | --- | --- | --- | --- | --- |
| NPV val | 0.990 | 0.980 | 0.975 | 0.950 | 0.900 | 0.850 |
| NPV | 0.973±0.021 | 0.979±0.021 | 0.971±0.022 | 0.929±0.034 | 0.867±0.041 | 0.849±0.031 |
| PPV | 0.318±0.096 | 0.299±0.109 | 0.369±0.156 | 0.523±0.161 | 0.789±0.173 | 1.000±0.000 |
| BACC | 0.748±0.023 | 0.746±0.030 | 0.775±0.032 | 0.748±0.041 | 0.596±0.063 | 0.527±0.014 |
| ACC | 0.629±0.086 | 0.609±0.101 | 0.681±0.105 | 0.822±0.085 | 0.859±0.033 | 0.850±0.030 |
| Sensitivity | 0.921±0.062 | 0.937±0.064 | 0.905±0.077 | 0.634±0.181 | 0.206±0.141 | 0.055±0.029 |
| Specificity | 0.575±0.107 | 0.554±0.121 | 0.644±0.136 | 0.862±0.127 | 0.985±0.016 | 1.000±0.000 |
| F1 | 0.460±0.091 | 0.439±0.107 | 0.498±0.125 | 0.536±0.097 | 0.290±0.155 | 0.103±0.053 |
| Threshold | 0.146±0.070 | 0.151±0.062 | 0.169±0.067 | 0.332±0.106 | 0.592±0.072 | 0.793±0.089 |

**Table 2:** Hyperparameters for grid search.

| Model | Hyperparameters |
| --- | --- |
| SNN | *lr*: {1e-3, 2e-4, 1e-4}, *n_val_stops*: {20}, *weight_decay*: {1e-5},<br>*intermediate_size*: {4, 16, 64}, *n_layers*: {1,3,6}, *alpha_dropout*: 0, 0.9, *optimizer*: {Adam} |
| KNN | *n_neighbors*: {3,11,25,51,101,201,301}, *weights*: {uniform, distance} |
| LR | *lr*: {1e-2, 1e-3, 5e-4, 1e-4}, *n_val_stops*: {20}, *weight_decay*: {1e-5}, *optimizer*: {Adam} |
| SVM (COVID-19) | *class*: {LinearSVC}, *dual*: {False}, *class_weight*: {None, balanced} |
| SVM (Mortality) | *class*: {SVC}, *kernel*: {linear, poly, rbf, sigmoid, precomputed}, *probability*: {True},<br>*class_weight*: {None, balanced} |
| RF | *n_estimators*: {501}, *criterion*: {gini, entropy}, *max_depth*: {2,8,32,None},<br>*min_samples_split*: {2}, *min_samples_leaf*: {1,8,32}, *max_features*: {auto, log2, None},<br>*max_leaf_nodes*: {None}, *class_weight*: {balanced, None} |
| XGB | *objective*: {binary:logistic}, *booster*: {gbtree, gblinear, dart}, *eta*: {0.1, 0.3, 0.6},<br>*gamma*: {0}, *max_depth*: {2,6,32}, *scale_pos_weight*: {True, False},<br>*grow_policy*: {depthwise, lossguide} |

**Table 3:** Comparison of estimated and actual performance. These metrics are calculated on the basis of the COVID-19 diagnosis prediction task with XGB. At significantly different deviations, the confidence intervals (CI) are colored in red. **a**, The actual performance is calculated on the listed month and the estimate is determined on the respective previous month. **b**, The estimate is determined by random samples from the 2020 cohort. **c**, The estimate is determined by random samples from the 2019 and 2020 cohort.

| **a** |  | Jun | Jul | Aug | Sep | Oct | Nov | Dec | Sum | Mean |
| --- | --- | --- | --- | --- | --- | --- | --- | --- | --- | --- |
|  | ROC AUC actual | 0.7994 | 0.7474 | 0.9225 | 0.8076 | 0.7766 | 0.7975 | 0.7419 | 0.4329 | 0.7990 |
|  | AUC estimate | 0.8182 | 0.7767 | 0.7880 | 0.9202 | 0.8302 | 0.7934 | 0.8219 |  | 0.8212 |
| | $\Delta$ ROC AUC actual - estimate | 0.0188 | 0.0293 | 0.1345 | 0.1126 | 0.0536 | 0.0042 | 0.0800 | | **0.0618** |
|  | CI95 lower ROC AUC actual | 0.6973 | 0.6687 | 0.8405 | 0.7014 | 0.7383 | 0.7789 | 0.7068 |  |  |
|  | CI95 upper ROC AUC actual | 0.8851 | 0.8262 | 0.9808 | 0.9014 | 0.8142 | 0.8179 | 0.7782 |  |  |
|  | CI95 lower ROC AUC estimate | 0.6983 | 0.6717 | 0.6959 | 0.8308 | 0.7276 | 0.7541 | 0.8040 |  |  |
|  | CI95 upper ROC AUC estimate | 0.9205 | 0.8684 | 0.8747 | 0.9878 | 0.9188 | 0.8297 | 0.8389 |  |  |
|  | PR AUC actual | 0.2413 | 0.1180 | 0.5155 | 0.3913 | 0.4540 | 0.7069 | 0.5878 | 0.9322 | 0.4307 |
|  | PR AUC estimate | 0.3123 | 0.2487 | 0.3948 | 0.6699 | 0.4678 | 0.5274 | 0.7256 |  | 0.4781 |
| | $\Delta$ PR AUC actual - estimate | 0.0711 | 0.1307 | 0.1207 | 0.2786 | 0.0138 | 0.1796 | 0.1378 | | **0.1332** |
|  | CI95 lower PR AUC actual | 0.1058 | 0.0704 | 0.2501 | 0.1967 | 0.3784 | 0.6714 | 0.5288 |  |  |
|  | CI95 upper PR AUC actual | 0.3941 | 0.1870 | 0.7190 | 0.5821 | 0.5325 | 0.7396 | 0.6428 |  |  |
|  | CI95 lower PR AUC estimate | 0.1442 | 0.1118 | 0.2397 | 0.4430 | 0.2768 | 0.4481 | 0.6916 |  |  |
|  | CI95 upper PR AUC estimate | 0.5236 | 0.4003 | 0.5448 | 0.8476 | 0.6356 | 0.6034 | 0.7617 |  |  |
| **b** |  | Jun | Jul | Aug | Sep | Oct | Nov | Dec | Sum | Mean |
|  | ROC AUC actual | 0.8116 | 0.7913 | 0.9531 | 0.8242 | 0.787 | 0.8227 | 0.7515 | 0.5034 | 0.8202 |
|  | ROC AUC estimate | 0.8959 | 0.8238 | 0.9075 | 0.8742 | 0.8922 | 0.8641 | 0.896 |  | 0.8791 |
| | $\Delta$ | 0.0843 | 0.0325 | 0.0456 | 0.0501 | 0.1052 | 0.0414 | 0.1444 | | **0.0719** |
|  | CI95 lower ROC AUC actual | 0.7096 | 0.6982 | 0.8955 | 0.7214 | 0.7500 | 0.8051 | 0.7156 |  |  |
|  | CI95 upper ROC AUC actual | 0.9074 | 0.8717 | 0.9889 | 0.9145 | 0.8237 | 0.8409 | 0.7888 |  |  |
|  | CI95 lower ROC AUC estimate | 0.8304 | 0.7229 | 0.8405 | 0.8072 | 0.8382 | 0.8151 | 0.8729 |  |  |
|  | CI95 upper ROC AUC estimate | 0.9598 | 0.9061 | 0.9589 | 0.9313 | 0.9394 | 0.9129 | 0.9185 |  |  |
|  | PR AUC actual | 0.2209 | 0.3593 | 0.6543 | 0.4538 | 0.5141 | 0.7191 | 0.6087 | 0.7654 | 0.5043 |
|  | P AUC estimate | 0.4047 | 0.4106 | 0.5428 | 0.3366 | 0.5122 | 0.5237 | 0.7129 |  | 0.4919 |
| | $\Delta$ | 0.1838 | 0.0513 | 0.1115 | 0.1172 | 0.002 | 0.1954 | 0.1042 | | **0.1093** |
|  | CI95 lower PR AUC actual | 0.1177 | 0.2050 | 0.4516 | 0.2654 | 0.4364 | 0.6856 | 0.545 |  |  |
|  | CI95 upper PR AUC actual | 0.4167 | 0.5227 | 0.8221 | 0.6239 | 0.5921 | 0.7522 | 0.6695 |  |  |
|  | CI95 lower PR AUC estimate | 0.2259 | 0.2431 | 0.3912 | 0.2057 | 0.3461 | 0.4121 | 0.6592 |  |  |
|  | CI95 upper PR AUC estimate | 0.6419 | 0.5858 | 0.7075 | 0.4794 | 0.6604 | 0.6353 | 0.7626 |  |  |
| **c** |  | Jun | Jul | Aug | Sep | Oct | Nov | Dec | Sum | Mean |
|  | ROC AUC actual | 0.7819 | 0.7638 | 0.9060 | 0.8483 | 0.7939 | 0.8226 | 0.7507 | 1.1024 | 0.8096 |
|  | ROC AUC estimate | 0.9900 | 0.9286 | 0.9820 | 0.9814 | 0.9623 | 0.9517 | 0.9734 |  | 0.9671 |
| | $\Delta$ | 0.2082 | 0.1647 | 0.0760 | 0.1332 | 0.1684 | 0.1291 | 0.2227 | | **0.1575** |
|  | CI95 lower ROC AUC actual | 0.6805 | 0.6603 | 0.7956 | 0.7448 | 0.7553 | 0.8048 | 0.7118 |  |  |
|  | CI95 upper ROC AUC actual | 0.8737 | 0.8567 | 0.9855 | 0.9338 | 0.8319 | 0.8412 | 0.7868 |  |  |
|  | CI95 lower ROC AUC estimate | 0.9800 | 0.8596 | 0.9656 | 0.9662 | 0.9197 | 0.9249 | 0.9648 |  |  |
|  | CI95 upper ROC AUC estimate | 0.9974 | 0.9807 | 0.9937 | 0.9929 | 0.9908 | 0.9746 | 0.9812 |  |  |
|  | PR AUC actual | 0.2274 | 0.3821 | 0.6260 | 0.4627 | 0.5326 | 0.7227 | 0.6073 | 1.3408 | 0.5087 |
|  | PR AUC estimate | 0.4554 | 0.4508 | 0.3643 | 0.4093 | 0.2814 | 0.2938 | 0.5585 |  | 0.4019 |
| | $\Delta$ | 0.2280 | 0.0688 | 0.2617 | 0.0535 | 0.2512 | 0.4289 | 0.0487 | | **0.1915** |
|  | CI95 lower PR AUC actual | 0.0973 | 0.2407 | 0.4050 | 0.2670 | 0.4548 | 0.6902 | 0.5489 |  |  |
|  | CI95 upper PR AUC actual | 0.3881 | 0.5292 | 0.8022 | 0.6256 | 0.6064 | 0.7565 | 0.6673 |  |  |
|  | CI95 lower PR AUC estimate | 0.2223 | 0.2635 | 0.2093 | 0.2563 | 0.1170 | 0.1982 | 0.4848 |  |  |
|  | CI95 upper PR AUC estimate | 0.6704 | 0.6217 | 0.5347 | 0.5634 | 0.4402 | 0.4298 | 0.6293 |  |  |

**Table 4:** Comparison of estimated and actual performance for mortality risk prediction. These metrics are calculated from the predictions of a RF, trained with the hyperparameters as determined in experiment (v). At significantly different deviations, the confidence intervals (CI) are colored in red. **a**, The actual performance is calculated on the listed month and the estimate was determined from the respective previous month. **b**, The estimate is determined by random samples from the positives cohort, occurring before the month, which the actual performance is calculated on.

| **a** | Sep | Oct | Nov | Dec | Sum | Mean |
| --- | --- | --- | --- | --- | --- | --- |
| ROC AUC actual | 0.8167 | 0.7427 | 0.7894 | 0.8403 | 0.2826 | 0.7973 |
| AUC estimate | 0.7279 | 0.9067 | 0.7716 | 0.8524 |  | 0.8146 |
| $\Delta$ ROC AUC actual - estimate | 0.0887 | 0.1640 | 0.0179 | 0.0121 | | **0.0707** |
| CI95 lower ROC AUC actual | 0.6462 | 0.6591 | 0.7525 | 0.7679 |  |  |
| CI95 upper ROC AUC actual | 0.9538 | 0.8146 | 0.8282 | 0.8980 |  |  |
| CI95 lower ROC AUC estimate | 0.3519 | 0.7619 | 0.6843 | 0.8156 |  |  |
| CI95 upper ROC AUC estimate | 1.0000 | 1.0000 | 0.8485 | 0.8888 |  |  |
| PR AUC actual | 0.3705 | 0.2645 | 0.2967 | 0.5548 | 0.7174 | 0.3716 |
| PR AUC estimate | 0.5233 | 0.6694 | 0.4188 | 0.5172 |  | 0.5322 |
| $\Delta$ PR AUC actual - estimate | 0.1528 | 0.4049 | 0.1221 | 0.0376 | | **0.1793** |
| CI95 lower PR AUC actual | 0.1329 | 0.1791 | 0.2321 | 0.3846 |  |  |
| CI95 upper PR AUC actual | 0.8302 | 0.3642 | 0.3724 | 0.7012 |  |  |
| CI95 lower PR AUC estimate | 0.0556 | 0.2417 | 0.2591 | 0.4242 |  |  |
| CI95 upper PR AUC estimate | 1.0000 | 1.0000 | 0.6097 | 0.6256 |  |  |
| **b** | Sep | Oct | Nov | Dec | Sum | Mean |
| ROC AUC actual | 0.8500 | 0.7714 | 0.8341 | 0.8686 | 0.2995 | 0.8310 |
| AUC estimate | 0.9063 | 0.9647 | 0.8564 | 0.8962 |  | 0.9059 |
| $\Delta$ ROC AUC actual - estimate | 0.0563 | 0.1933 | 0.0223 | 0.0276 | | **0.0749** |
| CI95 lower ROC AUC actual | 0.7071 | 0.6757 | 0.7915 | 0.8079 |  |  |
| CI95 upper ROC AUC actual | 0.9692 | 0.8463 | 0.8722 | 0.9155 |  |  |
| CI95 lower ROC AUC estimate | 0.8031 | 0.9050 | 0.7514 | 0.8448 |  |  |
| CI95 upper ROC AUC estimate | 0.9824 | 1.0000 | 0.9477 | 0.9428 |  |  |
| PR AUC actual | 0.5203 | 0.3519 | 0.4746 | 0.5772 | 1.2041 | 0.4810 |
| PR AUC estimate | 0.7729 | 0.9367 | 0.702 | 0.7164 |  | 0.7820 |
| $\Delta$ PR AUC actual - estimate | 0.2526 | 0.5848 | 0.2274 | 0.1393 | | **0.3010** |
| CI95 lower PR AUC actual | 0.1694 | 0.2241 | 0.3763 | 0.4277 |  |  |
| CI95 upper PR AUC actual | 0.8972 | 0.5302 | 0.5750 | 0.7167 |  |  |
| CI95 lower PR AUC estimate | 0.5043 | 0.7980 | 0.4771 | 0.5783 |  |  |
| CI95 upper PR AUC estimate | 0.9455 | 1.0000 | 0.8756 | 0.8256 |  |  |

**Table 5:**
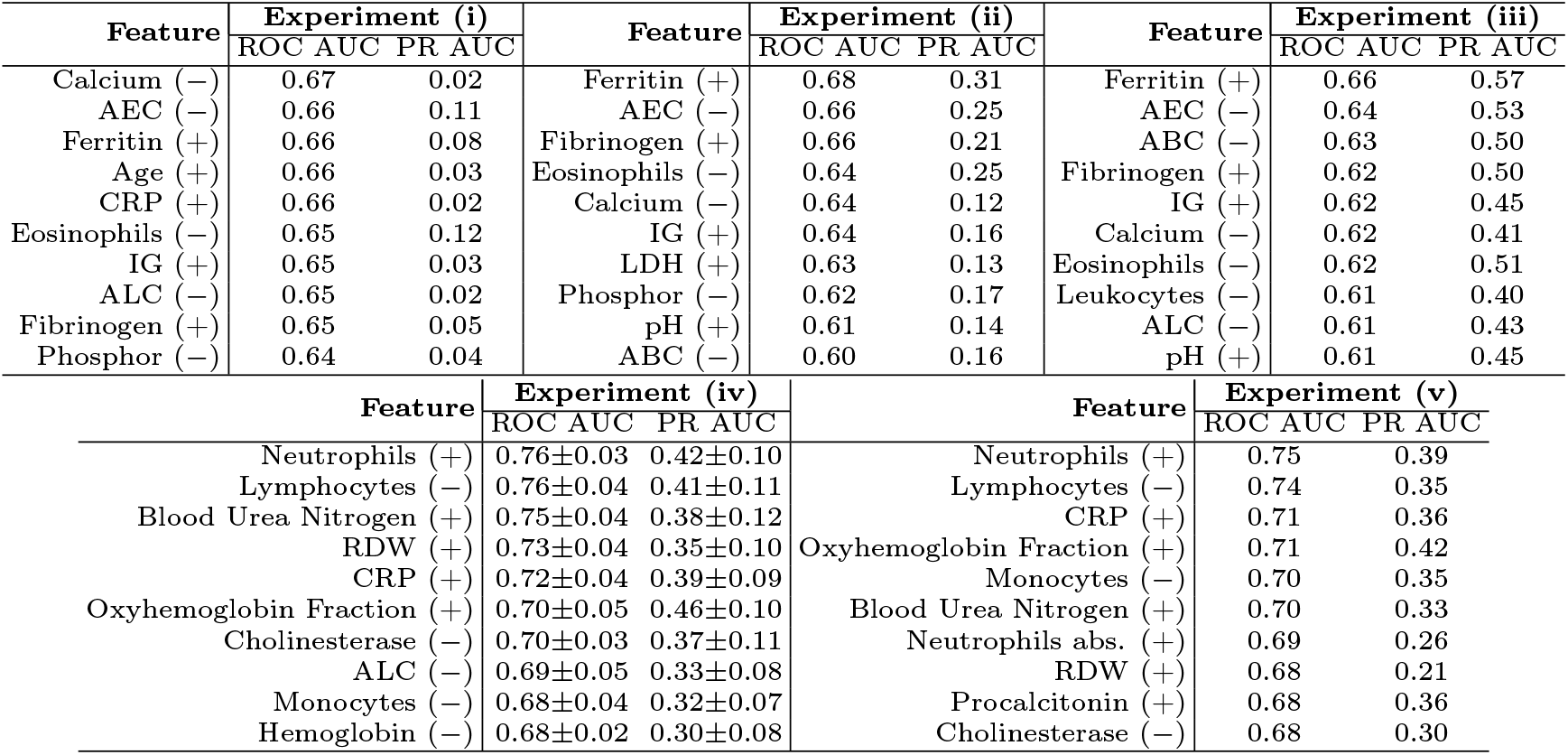
Features with discriminating capability. Top-10 features as predictors for COVID-19 diagnosis (experiment (i)-(iii)) and mortality prediction (experiment (iv) and (v)). The sign in brackets indicates whether the target is connected with the positive (+) or the negative sign of the feature value (±), i.e., patients with high ferritin and low calcium have higher probability for class COVID-19 positive. The standard deviation (−) is listed for experiment (iv), for the other experiments the test set is fixed. Abbreviations: absolute eosinophil count (AEC), immature granulocytes (IG), absolute basophil count (ABC), lactate dehydrogenase (LDH), C-reactive protein (CRP), absolute lymphocyte count (ALC), red cell distribution width (RDW).

**Figure 3:**
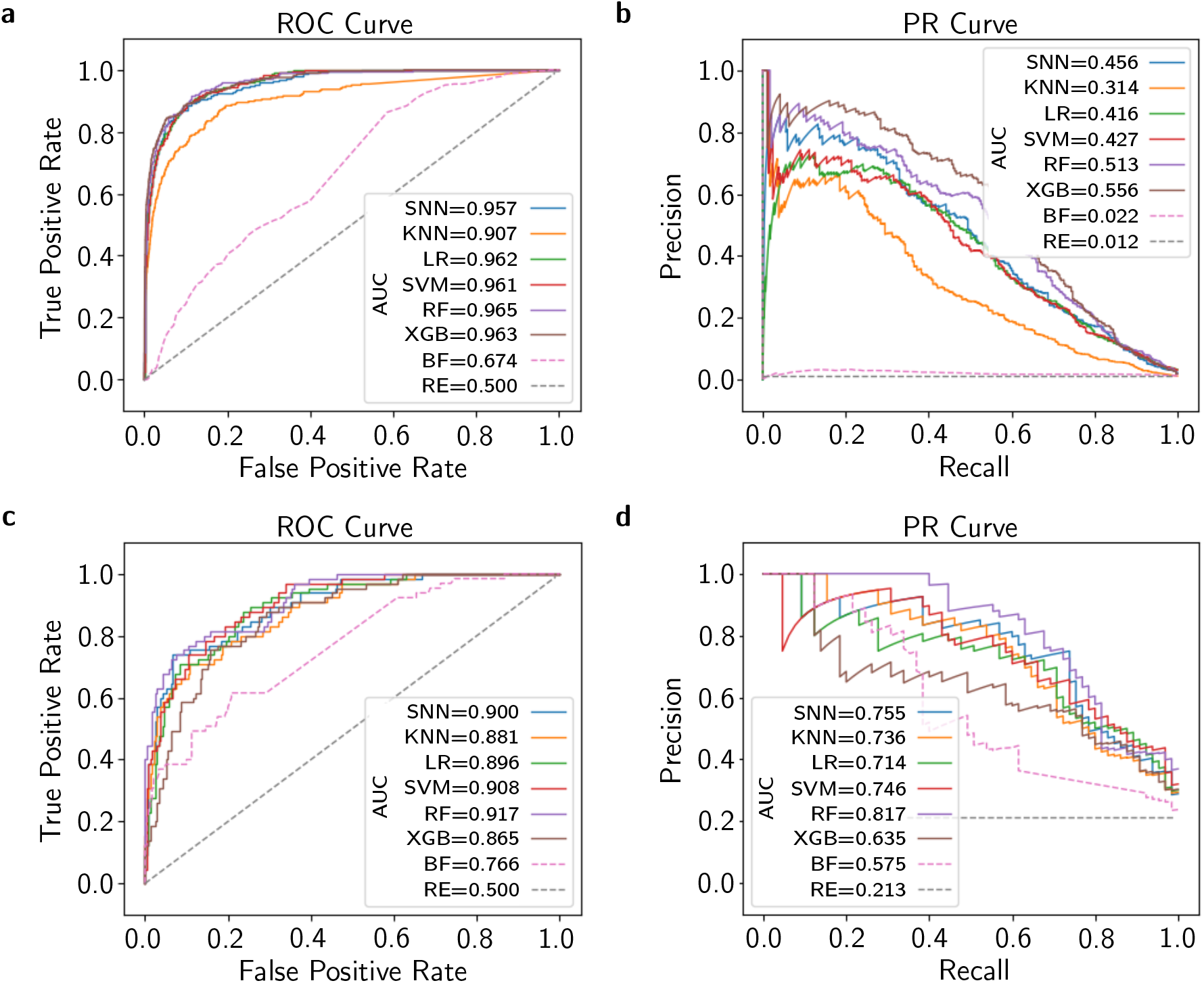
Comparison of model classes for COVID-19 diagnosis and mortality prediction. **a**, ROC and **b**, PR curves for the test set of COVID-19 diagnosis prediction in experiment (i). **c**, ROC and **d**, PR curves for mortality prediction in experiment (iv). **a**-**d**, Curves plotted for the different model classes at one random seed. RF and XGB outperform the other model classes as well as the random estimator (RE) baseline and the best feature (BF) as an estimator.

#### (i) COVID-19 diagnosis prediction & random cross-validation with pre-pandemic negatives

In this experiment all cases from 2019 and 2020 are randomly shuffled, see Section 4.3 for more details. In experiment (i) the highest performance is achieved, however, domain shifts are not considered in the performance estimate. The model with the highest ROC AUC of 0.97±0.00 and PR AUC of 0.52±0.01 is the RF. Note that the baseline of a random estimator (RE) is at 0.50±0.00 for ROC AUC and 0.01±0.00 for PR AUC, the latter due to the high imbalance of positive and negative samples. For in-hospital application a threshold is required to classify the probabilities of the models to the positive or negative class. This threshold is a trade-off between identifying all positive cases and a low number of false positives. Therefore, we report the thresh-old-dependent metrics for multiple thresholds, which are determined by defining negative predictive values on the validation set. The results with these determined thresholds are shown in Table 1 **c** for the RF.

#### (ii) COVID-19 diagnosis prediction & random cross-validation with recent negatives

The test set of experiment (ii) only comprises cases, which have been tested for COVID-19 with an RT-PCR test. The 2020 cohort comprises patients which are suspicious for COVID-19, some might even have characteristic symptoms. Therefore, a classification of the samples in the 2020 cohort is more difficult and potential biases between the 2019 and 2020 cohort cannot be exploited. XGB outperforms the other models with a ROC AUC of 0.92±0.00 and a PR AUC of 0.62±0.00.

#### (iii) COVID-19 diagnosis prediction & temporal cross-validation

In experiment (iii), the model is trained with samples until October and evaluated on samples from November and December. XGB achieves the highest ROC AUC of 0.81±0.00 and a PR AUC of 0.71±0.00. We face an additional performance drop in comparison to experiment (i) and (ii), which points to a domain shift over time. Besides others, this domain shift over time occurs due to potential changes in the lab infrastructure, testing strategy, prevalence of COVID-19 in different patient groups, or maybe even due to mutations of the COVID-19 virus, see Figure 1 and Section 1 for more details. These results again emphasize the necessity for countering the domain shifts with lifelong learning and assessment.

#### (iv) Mortality prediction & random cross-validation

We predict the mortality risk of COVID-19 positive patients, who only occur in the 2020 cohort. The samples are randomly shuffled and a five-fold nested cross-validation is performed. RF outperforms the other models for the mortality prediction with a ROC AUC of 0.88±0.02 in (iv) and a PR AUC of 0.63±0.11. We report the threshold-dependent metrics in Table 1 **d**, although the prediction scores of survival or death, provided by our models, are more informative for clinicians in practice, rather than a hard separation by a threshold into the two classes.

#### (v) Mortality prediction & temporal cross-validation

In experiment (v), the model is trained with samples until October and evaluated on samples from November and December for mortality prediction of COVID-19 positive patients. Again, RF outperforms the other models with a ROC AUC of 0.86±0.01 and a PR AUC of 0.56±0.01. The performance drops from experiment (iv) to (v), revealing a domain shift over time for mortality prediction.

### 2.5 Lifelong learning and assessment

We propose re-training and re-assessment with high frequencies to tackle the domain shifts by exploiting the new samples to achieve high performance and model credibility in the real world hospital setting. Therefore, we suggest to continuously determine the COVID-19 status with an RT-PCR test of some patients to acquire frequent samples, which is indispensable to avoid the model behavior to drift into unexpected and poor performance. These measures are essential to enable trustworthy ML models for clinical utility.

The effect of the re-training frequency of the model is shown in Figure 4 **b**. The performance of the ML models increases with the re-training frequency, thereby reducing the domain shift of the training to the test samples. The evaluation procedure is shown in Figure 4 **a** and in Section 4.4.

**Figure 4:**
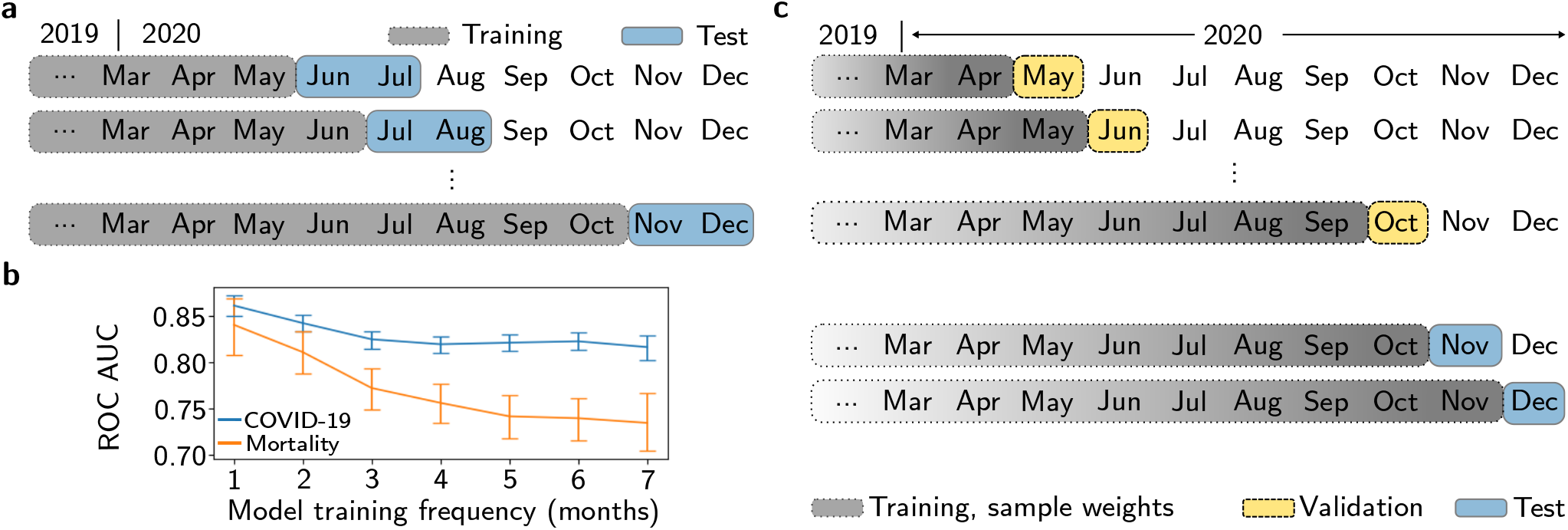
Lifelong learning. **a**, Evaluation for a model training frequency of two months. The model is evaluated with an intercept of one month, but the model is evaluated on the two subsequent months after training. **b**, Effect of model training frequency on performance. The mean and the 95 % confidence intervals (error bars) of the ROC AUCs. The ROC AUC performance decreases with lower model training frequency. **c**, Current samples are weighted higher in training to counter the domain shifts. The weighting is selected on the validation months starting from May until October. The selected weighting is evaluated on the test months November and December.

To counter the domain shifts, we additionally propose to weight current samples stronger during training of the COVID-19 diagnosis model, see Figure 4 **c**. On the validation set (May - October), we determine the best weighting in dependence of the sample recency. The highest performance gain on the validation set is achieved by setting the weight of the 2019 cohort samples to 0.01 and the weight of the samples of the most recent month to 3, and the second last month to 2 ([1, 1, 2, 3]). Compared to weighting all samples equally, this increases the ROC AUC on the validation set from 0.8118 (95 % CI: 0.7849-0.8386) to 0.8502 (95 % CI: 0.8271-0.8734) (p-value = 9e-6), which is statistically significant. The selected weighting is tested on November and December, leading to a statistically significant increase of the ROC AUC from 0.7996 (95 % CI: 0.7831-0.8162) to 0.8120 (95 % CI: 0.796-0.828) (p-value =0.0045). The method to determine the weighting is described in more detail in Section 4.4.

### 2.6 Features with discriminating capability

For clinical insight, the violin plots show discriminating capability of the selected features for the three different cohorts (2019 and 2020 cohort, 2020 cohort, COVID-19 positive cohort) in Figure 5. The plotted features are selected based on their ROC AUC on the five experiments, the top-10 features as predictors for all five tasks are listed in Table 5 in the supplementary material.

**Figure 5:**
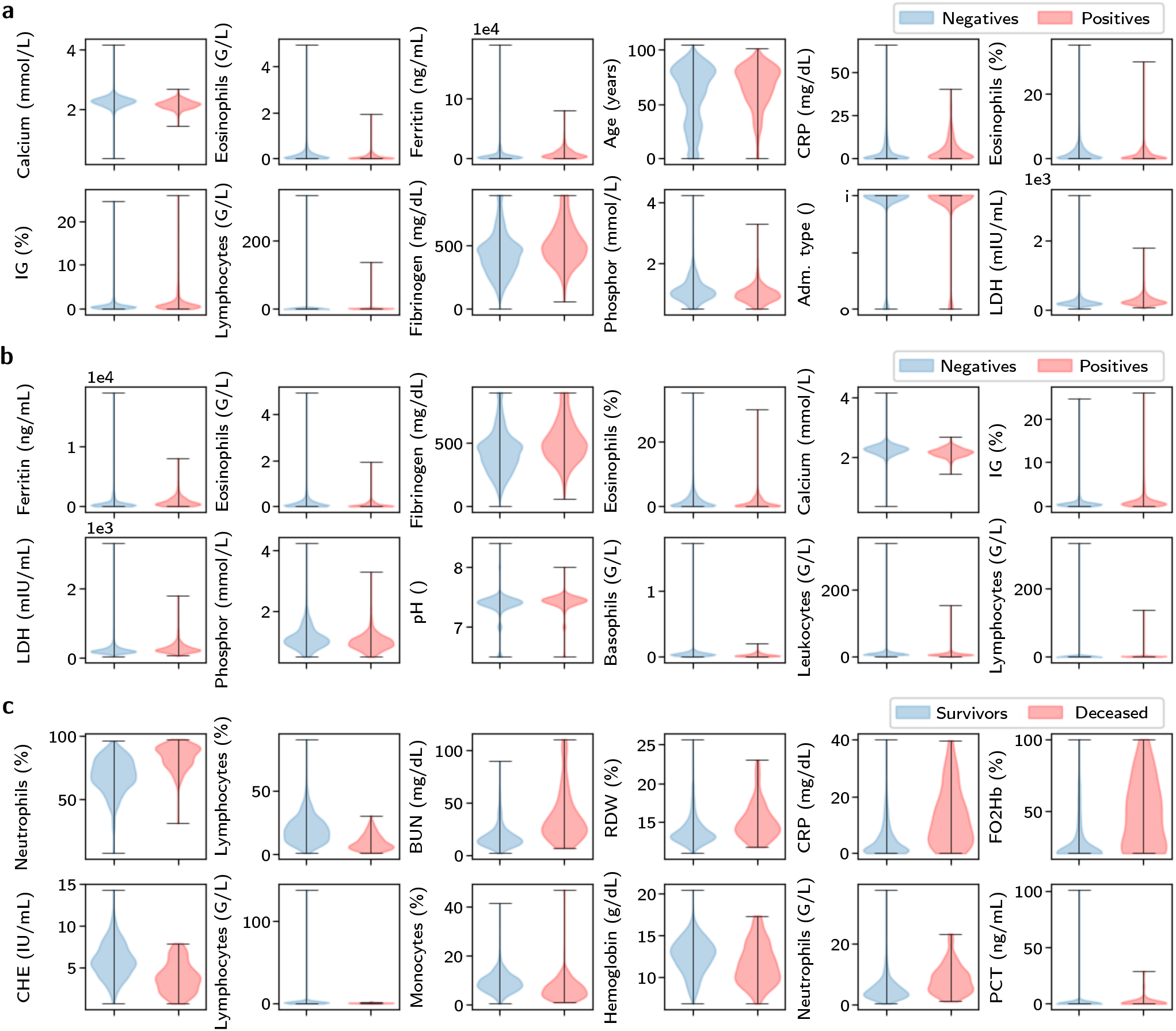
Features with discriminating capability. **a**, Measured features in the 2019 and 2020 cohort for COVID-19 diagnosis prediction for negative and positive class. **b**, Features with discriminating capability for COVID-19 diagnosis prediction in the 2020 cohort, which contains the RT-PCR tested patients. **c**, Measured features of positives cohort for mortality prediction for survivors and deceased. Abbreviations: C-reactive protein (CRP), immature granulocytes (IG), type of hospital admission (Adm. type), inpatient (i), outpatient (o), lactate dehydrogenase (LDH), pH-value (pH), blood urea nitrogen (BUN), red cell distribution width (RDW), oxyhemoglobin fraction (FO2Hb), cholinesterase (CHE), procalcitonin (PCT).

**Figure 6:**
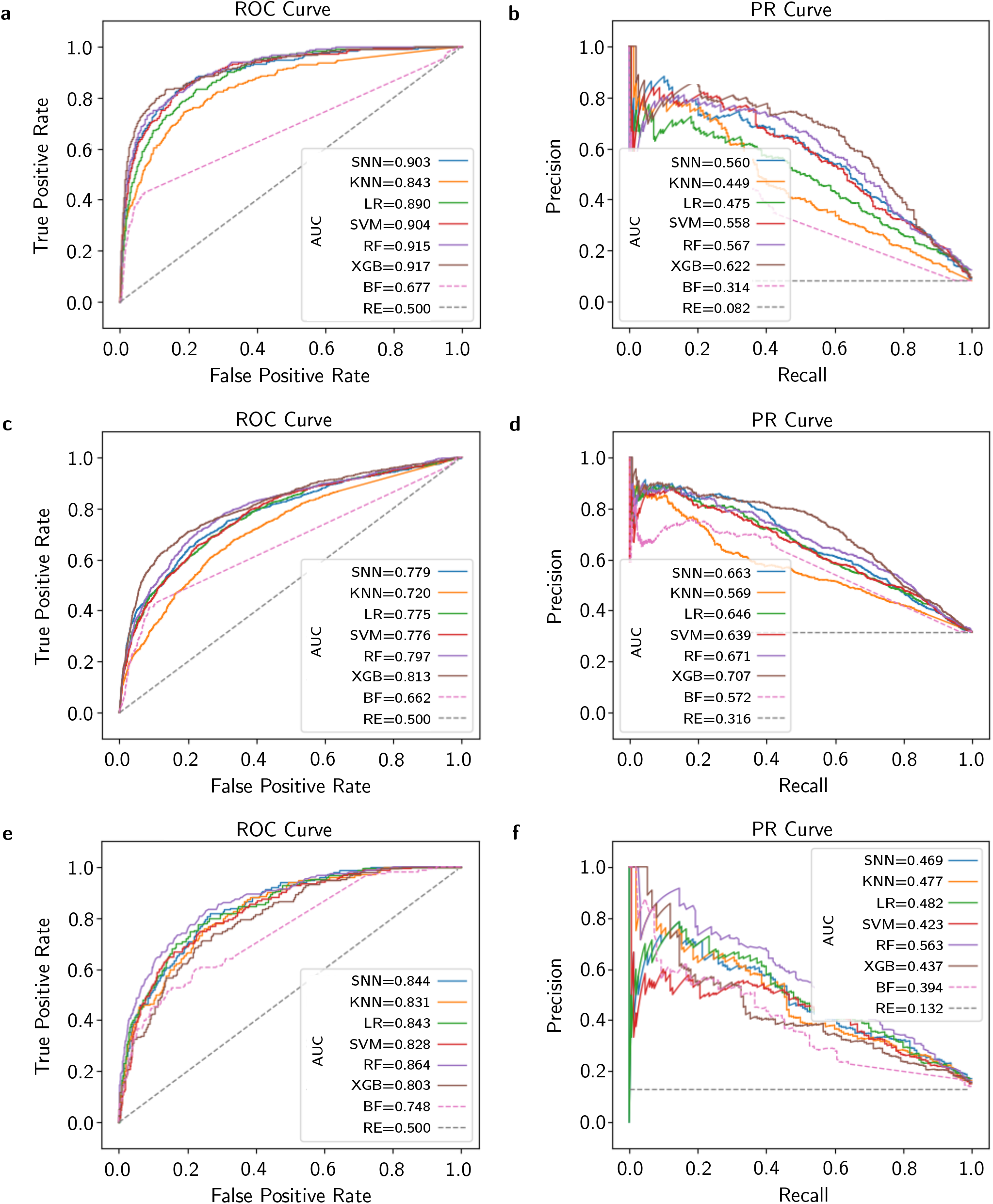
Comparison of ML models for different experiments. ROC and PR curves for test set of **a** and **b**, experiment (ii); **c** and **d**, experiment (iii); **e** and **f**, experiment (v); the curves are plotted for different models (one random seed) and compared with best feature (BF) and random estimator (RE).

**Figure 7:**
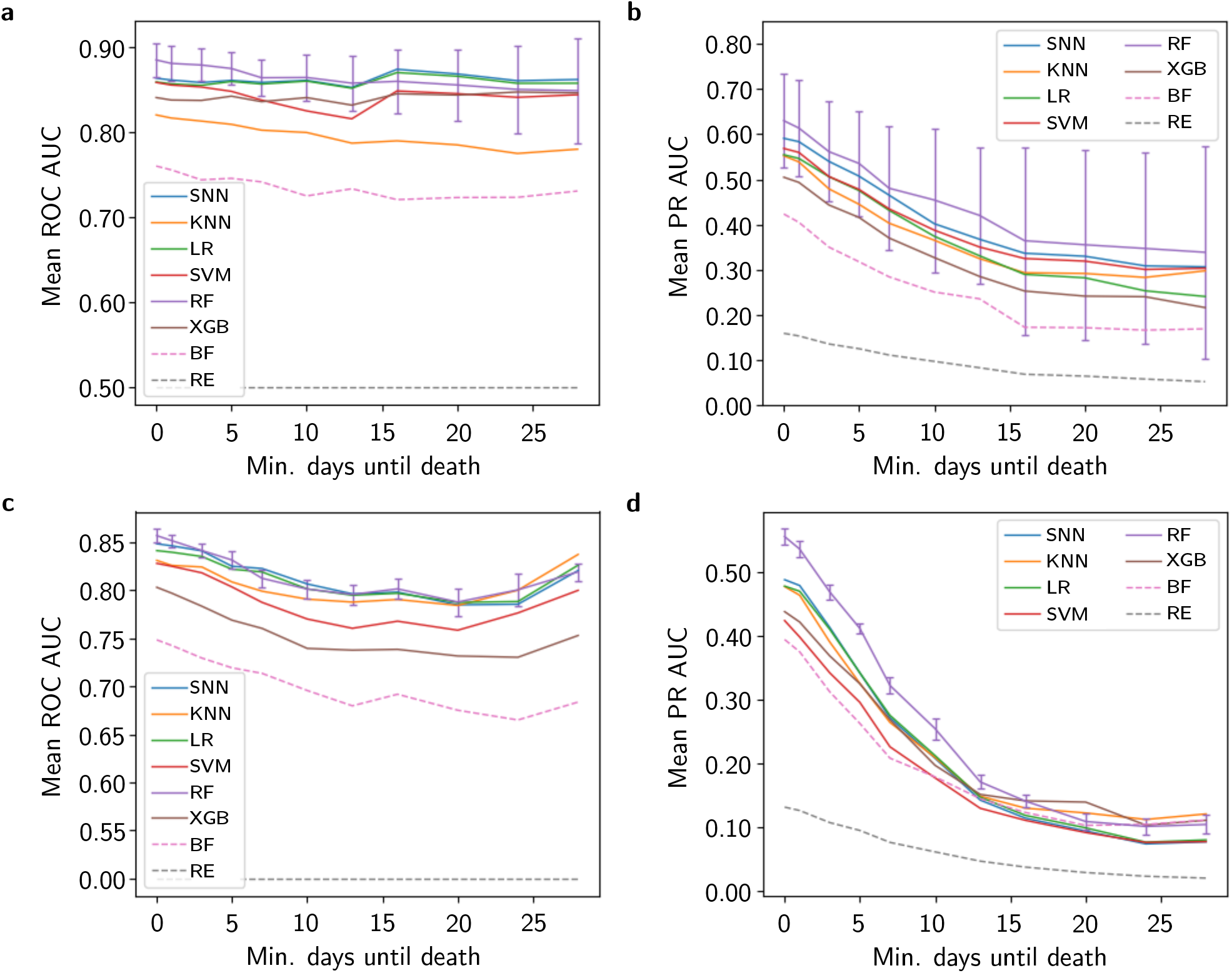
Performance of models in dependence of the minimum number of days until death. In this figure we answer, how early before death we can predict the risk of dying. Samples at which the death has occurred within the next days (minimum days until death), are excluded from the test set (but not from the training and validation set). **a**, The mean of ROC AUC and **b**, PR AUC values of five test folds is plotted. **c**, The mean of ROC AUC and **d**, PR AUC values of five random seeds in prospective evaluation are shown. For visual clarity, the standard deviations (error bars) are only plotted for the RF. The mortality risk can be estimated early before death, as the discriminating capability of the models remains high with increasing number of minimum days until death. The mean PR AUC in **b** and **d** is decreasing with increasing minimum days until death, equally to the random estimator baseline, due to the decreasing ratio of deceased to survivors.

**Figure 8:**
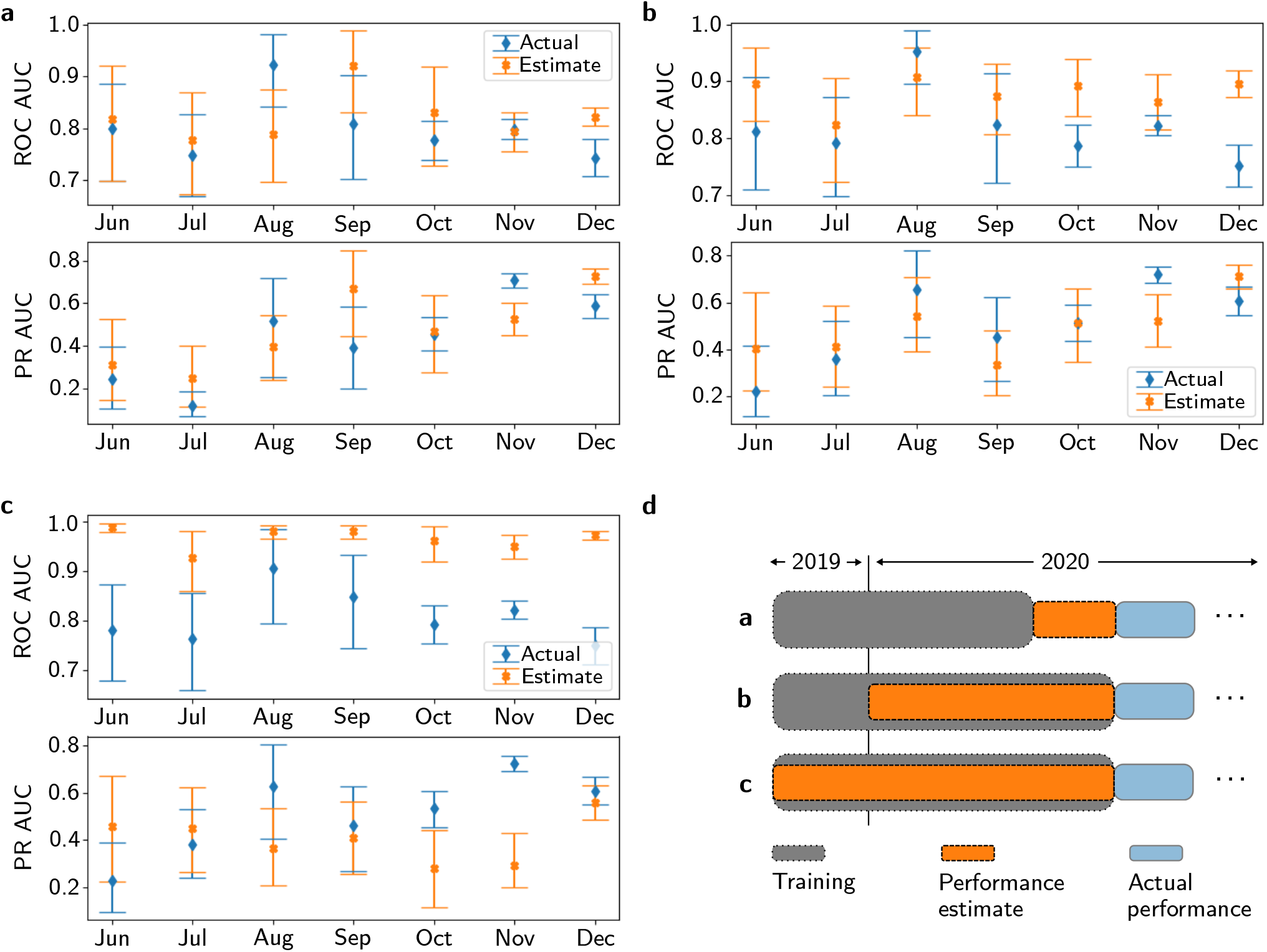
Deviation of estimated from actual performance with three options to determine the performance estimate. **a**, The actual ROC AUC differs significantly from the estimate in December, the PR AUC in November and December. **b**, Significant difference in October and November for ROC AUC and in November for PR AUC. **c**, Estimated and actual ROC AUC are significantly different in all month but August, due to heavy domain shifts, and PR AUC in October and November. The mean deviation of estimated and actual ROC AUC and PR AUC is higher in **c** compared to **a** and **b. d**, Three options to determine the performance estimate. In **a**, the performance estimate is calculated from the preceding month. In **b**, the samples for the estimate are randomly selected from the 2020 cohort (20 %), and in **c** they are randomly sampled from the 2019 and the 2020 cohort (20 % of the 2020 cohort and equal proportion from the 2019 cohort).

**Figure 9:**
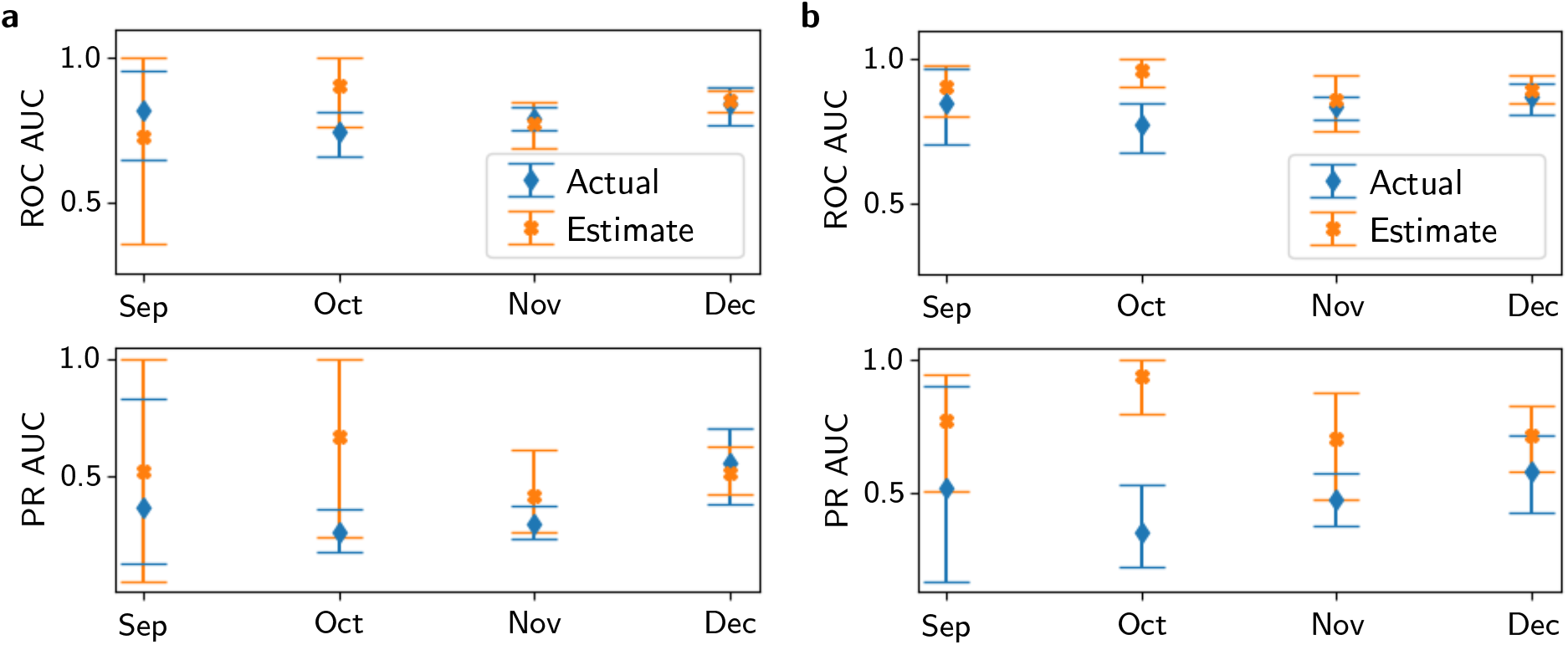
Deviation of estimated from actual performance for mortality risk with two options to determine the performance estimate. **a**, The estimate is determined on the respective previous month. Note that the confidence interval at an early stage of the pandemic is high due to a low number of samples. **b**, The estimate is determined on randomly sampled 20 % of the COVID-19 positives, who occurred before the actual performance month. There is a significant difference in October for ROC AUC and PR AUC, which means that the performance estimate is overoptimistic in October.

## 3 Discussion

Through multiple experiments we expose domain shifts and their detrimental effect on ML models for COVID-19 diagnosis. We suggest to carefully assess the model performance frequently to avoid unexpected behavior with potentially adverse consequences, such as even greater spread of the disease due to trusting the wrongly classifying model. The model should be re-trained after particular time-periods to exploit newly acquired samples and, thus, to countermand the domain shift effect. To this end, we propose to assign a higher weight to recent samples, which, as we show, increases the predictive performance.

In this large-scale study, we train and evaluate our models with more samples than most studies^18–22^. Besides our large number of tested subjects, we also exploit pre-pandemic negative samples, which vastly increases our data set size. In comparison to Soltan et al.^23^ and Plante et al.^24^ we use the pre-pandemic as well as the pandemic negatives in our data set.

We achieve high predictive performance with our models, comparable to previous studies^18,19,21,25,35^, although the results can not be directly compared as our assessment procedure is more rigorous. Different assessment procedures within our study also yield highly variable performance estimates. Some studies suggested logistic regression models for COVID-19 and mortality prediction^39,53^, however, most identified (X)GB or RF as the best model classes^18,20,25,31,38^. We confirm these findings and suggest to use XGB or RF for COVID-19 diagnosis and RF for mortality prediction, as these exhibit the highest performance in our experiments.

Our models only require a small set of features of a patient, concretely a minimum of twelve blood test parameters and the age, gender and hospital admission type; in total at least 15 features. In case many blood test parameters are available, the model exploits up to 100 pre-selected features. Missing features are imputed, thereby allowing model application also on samples with a small number of features. This enables automatically scanning the blood tests without additional effort by the hospital staff, as opposed to published models, which require more expensive features, e.g., vital signs, which might not be as easily available^19,22,23^.

One limitation of our work could be that we did not evaluate the generalization of our model to other hospitals. A transfer of a COVID-19 diagnostic model should only be done with thorough re-assessments, as a domain shift between hospitals might be present. Besides others, such domain shifts from one institution to another could result from different testing strategies, laboratory equipment or demographics of the population in the hospital catchment area. Re-training of models rather than transferring to another hospital should be considered to obtain a skilled and trustworthy model. However, this is not part of our investigation. Our findings and suggestions about domain shifts should be accounted for in all hospitals when applying a COVID-19 model.

We evaluate our models on different cohorts to show the high performance as well as to reveal the domain shifts. However, the 2020 cohort only contains subjects that were tested for COVID-19 and where a blood test was taken. Hence, the 2020 cohort only is a subset of the total patient cohort on which the model will be applied. To counteract missing samples from a particular group, we also use the pre-pandemic negatives, which should cover a wide variety of negatives due to the large data set. An evaluation of all blood tests of 2020 just is not possible due to the lack of RT-PCR tests which serve as labels in our ML approach. Non-tested subjects of 2020 cannot be assumed to be negatives, therefore we discard them. This could only be circumvented by explicitly testing a large number of patients for this study, who would not be tested otherwise.

For lifelong learning and assessment, testing of a subset of the patients with an RT-PCR test still is necessary to identify and counter the temporal domain shift. However, this does not diminish the benefit of the model as by automatically scanning all blood tests a large number of patients can be checked for COVID-19, which would not be feasible with expensive and slow RT-PCR tests. The benefit of the model also transfers to hospitals or areas with limited testing capacity. Rather than replacing RT-PCR tests, the model can be applied as a complement to or replacement for antigen tests. The model can be retrained with the already implemented pipeline. The computational effort is relatively low, as the model only requires tabular data and no time series (sound recordings) or images (CT, X-ray)^11–17^. Other studies do not consider the domain shifts and the associated necessity for re-training, although it is indispensable for clinical utility. Lifelong learning and assessment does not only provide a performance gain for diagnostic models in pandemics like COVID-19, but also for other medical tasks, or in general, other applications of ML, where we face a continuous stream of data.

We demonstrate the high capability of ML models in detecting COVID-19 infections and COVID-19-associated mortality risk on the basis of blood tests on a large-scale data set. With our findings concerning domain shifts and lifelong learning and assessment, we want to advance the ML models to be accurate and trustworthy in real world hospital settings. Lifelong learning and assessment is an important tool to allow the transition of research results to actual application in hospitals. By advancing this field of research, we want to increase patient safety and protect clinical staff and we wish to make a contribution in banning the pandemic.

## 4 Methods

### 4.1 Ethics approval

Ethics approval for this study was obtained from the ethics committee of the Johannes Kepler University, Linz (approval number: 1104/2020). The study is conducted on the blood tests (including age, gender and hospital admission type) from July 2019 until December 2020 and the COVID-19 RT-PCR tests from 2020 of the Kepler University Hospital, Med Campus III, Linz, Austria. In our study, we analyze anonymized data only.

### 4.2 Data set preparation

We predict the result of the RT-PCR COVID-19 test on the basis of routinely acquired blood tests. A block diagram of the data set is sketched in Figure 2. We only use the blood tests before the RT-PCR test to avoid bias caused by the test result. We limit the time deviation of the blood test to the COVID-19 RT-PCR test to 48 hours to ensure that the blood test matches the determined COVID-19 status. Additionally, we incorporate pre-pandemic blood tests from the year 2019 as negatives to our data set to cover a wide variety of COVID-19 negative blood tests. For the data from the year 2020, we aggregate the blood tests of the last 48 hours before the test. If parameters are measured more than once, we take the most recent one, see Figure 2 **b**. In case no COVID-19 test follows the blood test within 48 hours, the blood test samples are discarded. Additionally, we discard all samples with a deviating RT-PCR test result within the next 48 hours, as the label might be incorrect. The data from 2019 does not contain COVID-19 tests, therefore, blood tests with a temporal distance of less than 48 hours are aggregated. The features age, gender and admission type (inpatient or outpatient) are added to the samples. For the prediction of the COVID-19 diagnosis, we select the 100 most frequent features in the 2019 cohort as the feature set. For the mortality task the 100 most frequent features are selected based on the positives cohort, as the model is only applied to COVID-19 positive samples. Each sample requires a minimum of 15 features (minimum of any twelve blood test features and age, gender and hospital admission type). All other features and samples are discarded. The fact that the samples only require a minimum of 15 features can lead to many missing entries as the feature vector has a length of 100. For each sample we create 100 additional binary entries, which indicate whether each of the features is missing or measured. The missing values are filled by median imputation. Hence, the models can be applied to blood tests with few measured values.

### 4.3 Experiments for model performance under domain shift

Given the presence of domain shifts, we define five experimental designs to estimate the performance. The experiments differ at the data split into training, validation and test set. These splits are conducted on patient level, such that one patient only occurs in one of the sets. In the first three experiments we train models for COVID-19 diagnosis prediction. We train and evaluate the COVID-19 diagnosis models with five random seeds with a fixed data split.

i. In our first experiment we randomly shuffle all patients and we split regardless of the patient cohorts (60 % training, 20 % validation, 20 % testing).
ii. The training and validation sets include the 2019 cohort and 80 % (60 % training, 20 % validation) of the 2020 cohort. The test set comprises the remaining samples (20 %) of the 2020 cohort. Therefore, the performance is estimated on patients who actually were tested for COVID-19.
iii. The training and validation sets include the 2019 cohort and the 2020 cohort before November (80 % training, 20 % validation). We conduct a prospective performance estimate for the test set with all samples from November and December 2020. In experiment (iv) and (v) we train the models to predict the mortality risk of COVID-19 positive patients.
iv. The training (60 %), validation (20 %) and test (20 %) sets comprise the positive cases from the 2020 cohort. Due to the limited number of samples, we predict performance with five-fold nested cross validation.
v. The training and validation sets include the positive cases from 2020 before November (80 % training, 20 % validation). The test set comprises the cases from November and December. The test set is fixed, but again, we train and evaluate the models with five random seeds.

Z-score normalization is applied to the entire data set, with the mean and standard deviation calculated from the respective training set.

We compare multiple different models suitable for tabular data. The pre-processing, training and evaluation is implemented in Python 3.8.3. In particular, the model classes RF, KNN and SVM are trained with the scikit-learn package 0.22.1. XGB is trained with the XGBClassifier from the Python package XGBoost 1.3.1. The SNN and LR are trained with Pytorch 1.5.0.

The models are selected and evaluated based on the ROC AUC^56^, which is a measure of the model’s discriminating power between the two classes. Further, we report the PR AUC^56^ and we calculate threshold-dependent metrics, where the classes are separated into positives and negatives, instead of probability estimates. These metrics are negative predictive value (NPV), positive predictive value (PPV), balanced accuracy (BACC), accuracy (ACC), sensitivity, specificity and the F1-score (F1)^57^. We additionally report the thresholds, which are determined on the validation set to achieve the intended NPV.

We perform a grid search over hyperparameters of the models, see Table 2 in the supplementary material. The best hyperparameters are selected based on the ROC AUC on the validation set. In the COVID-19 diagnosis prediction tasks (experiment (i)-(iii)) we use one fixed validation fold due to the high number of samples. The models are trained and evaluated with five random seeds. For the mortality prediction tasks (experiment (iv) and (v)) the mean ROC AUC over five validation folds is calculated to select the hyperparameters. Further, the selected models are evaluated on the test set to estimate the performance. Experiment (iv) is evaluated with five-fold nested cross validation and all other experiments use a fixed test set. The mean and standard deviation of the models, which are trained, validated and tested with five random seeds, are reported.

### 4.4 Experiments for lifelong learning and assessment

We conduct three experiments to show the necessity of lifelong learning and assessment for trustworthy and accurate models. The first experiment investigates the deviation of the estimated to the actual performance. Therefore, we test the models on the months June until December. The performance estimate is calculated on the respective preceding month (May until November), see Figure 1 **b**. The 95 % confidence intervals are determined via bootstrapping by sampling 1,000 times with replacement. The deviations of estimated and actual performance are checked for significance. For this purpose, XGB is trained with the selected hyperparameters of experiment (iii).

Further, we check the effect of the model training frequency on the performance. We evaluate the trained model on different numbers of subsequent months without re-training. We also refer to this number of subsequent months as model training frequency. A model training frequency of two months is sketched in Figure 4 **a**. We evaluate the different model training frequencies with an increment of one month, concatenate the predictions as well as the targets and calculate the ROC AUC and its 95 % confidence interval with bootstrapping 1,000 times with replacement. We do not report PR AUC, as the prevalence in the test sets of the different model training frequencies are not comparable.

In our third experiment for lifelong learning and assessment, we investigate the effect of higher weights for current samples during training, as shown in Figure 4 **c**. Therefore, we define May until October as our validation months to select the optimal weighting and we evaluate the selection on November and December. We train the models with all available data before the respective validation month with the best hyper-parameters determined in experiment (iii). The predictions and targets are concatenated for all validation months. With a one-sided, paired DeLong test^58^, we test our hypothesis that the ROC AUC increases when current samples are weighted higher than older samples, in comparison to the ROC AUC when all samples are equally weighted. We pass the concatenated prediction and target vectors to the DeLong test, which returns the p-value and ROC AUC, calculated with the pROC package 1.17.0.1 in R.

We identify the best weighting by combining all listed options of weights of the 2019 cohort and of the most recent, previous months on the validation set. The default weight of the samples is 1. We restrict the 2019 cohort weights to the set: {1, 0.1, 0.01, 0.001 }, and the weights of the previous months to: { [1, 1, 1, 1], [1, 1, 1, 2], [1, 1, 2, 3], [1, 2, 3, 4], [2, 3, 4, 5] }, with the last entry in each square bracket being the weight of the last month, the second last of the second last month, and so forth. Afterwards, we normalize the weights to the length of the training samples, thereby we only change the relative weighting. As determined by the hyperparameter search, we also pass the scaling factor to `scale_pos_weight` to the model to balance positive and negative samples. The best weighting parameters are selected on the validation set and tested on November and December.

### 4.5 Features with discriminating capability

Besides the ML models, we additionally report statistical evaluations to allow clinical insight: we calculate the ROC AUCs of individual features equally to the five experiments in Section 4.3. For these evaluations, the features themselves are considered as predictors. This way, we can identify features with discriminating capability and compare these with the ML models. The ROC AUC is equivalent to the concordance-statistic (c-statistic) for binary outcomes^59^. Note that we do not train a model for this purpose, we simply use the positive or negative feature value as a predictor on the test set. Thereby, we identify important features for the COVID-19 diagnosis and the mortality task (Table 5 in supplementary material). Additionally, we visually prepare the most important features selected from the above described evaluation. The features of the full data set (2019 and 2020 cohort) and the 2020 cohort are plotted for the COVID-19 diagnosis as well as for the 2020 cohort for the mortality prediction in Figure 5. The violin plots only contain measured features, imputed feature values are not displayed for better visual clarity.

## Data Availability

The data set is not available for public use due to data privacy reasons.
Code is provided at https://github.com/ml-jku/covid.

## 5 Data availability

The data set is not available for public use due to data privacy reasons.

## 6 Code availability

Code is provided at https://github.com/ml-jku/covid.

## 8 Acknowledgements

We thank the projects Medical Cognitive Computing Center (MC3), AI-MOTION (LIT-2018-6-YOU-212), DeepToxGen (LIT-2017-3-YOU-003), AI-SNN (LIT-2018-6-YOU-214), DeepFlood (LIT-2019-8-YOU-213), PRIMAL (FFG-873979), S3AI (FFG-872172), DL for granular flow (FFG-871302), ELISE (H2020-ICT-2019-3 ID: 951847), AIDD (MSCA-ITN-2020 ID: 956832). We thank Janssen Pharmaceutica, UCB Biopharma SRL, Merck Healthcare KGaA, Audi.JKU Deep Learning Center, TGW LOGISTICS GROUP GMBH, Silicon Austria Labs (SAL), FILL Gesellschaft mbH, Anyline GmbH, Google, ZF Friedrichshafen AG, Robert Bosch GmbH, Software Competence Center Hagenberg GmbH, TÜV Austria, and the NVIDIA Corporation. We thank Franz Grandits, Innosol for the daily download of the age distribution data of the newly infected COVID-19 patients from BMSGPK.

## 9 Author contributions

T.R., T.T., J.M., S.H. and G.K. designed the study. C.B. exported and anonymized the data from the hospital information system. T.R., A.M. and T.T. preprocessed the blood tests. T.R. pre-processed the RT-PCR tests and mortality data. T.R. implemented the ML models and conducted the experiments. T.R., S.H. and G.K. wrote the manuscript. T.T. wrote the application for the ethics approval. S.H., J.M. and G.K. supervised the project. All authors critically revised the draft and approved the final manuscript.

## A Supplementary material

